# Internalizing and externalizing behaviors in school-aged children are related to state anxiety during magnetic resonance imaging

**DOI:** 10.1101/2021.08.11.21261892

**Authors:** Robin Eijlers, Elisabet Blok, Tonya White, Elisabeth M.W.J. Utens, Henning Tiemeier, Lonneke M. Staals, Johan M. Berghmans, Rene M.H. Wijnen, Manon H.J. Hillegers, Jeroen S. Legerstee, Bram Dierckx

**Author notes:** **Corresponding author** Tonya White, Department of Child and Adolescent Psychiatry/Psychology, Erasmus MC-Sophia / Kamer KP-2869, Postbus 2060, 3000 CB Rotterdam. These authors contributed equally (Robin Eijlers, Elisabet Blok). **Ethical standards** This study was approved by the Medical Ethical Committee of the Erasmus Medical Center and has been performed in accordance with the ethical standards laid down in the 1964 Declaration of Helsinki and its later amendments. All persons gave their informed consent prior to their inclusion in the study. **Declaration of Interest Statement** The authors declare no conflicts of interest. **Availability of data and material** Generation R data is available to researchers and requests should be directed to the management team of the Generation R Study. Release of data is contingent upon privacy and ethical restrictions.

## Abstract

Magnetic resonance imaging (MRI) procedures often evoke anxiety in children. Further, anxious children may be less likely to participate in MRI research, leading to a possible selection bias, and may be more likely to move during image acquisition, resulting in lower image quality and possible information bias. Therefore, state anxiety is problematic for functional and structural MRI studies. Children with behavioral problems, such as internalizing and externalizing behaviors, may be more likely to experience state anxiety prior to and during MRI scanning. Therefore, our first aim was to investigate the relationship between internalizing/externalizing behavior and children’s MRI-related state anxiety. Our second aim was to investigate the relationship between internalizing and externalizing behavior and MRI research participation. Our final aim was to investigate the effect of internalizing and externalizing behavior as well as MRI-related anxiety on image quality in children. We included 1,241 six- to ten-year-old children who underwent a mock MRI. Afterwards, if not too anxious, these children were scanned using a 3-Tesla GE Discovery MRI system (*n* = 1,070). Internalizing and externalizing behaviors were assessed with the Child Behavior Checklist. State anxiety was assessed with a visual analogue scale. Internalizing behaviors were positively associated with child state anxiety, as reported by child, parent, and researcher. For state anxiety reported by the parent and researcher, this relationship was independent of externalizing behaviors. Externalizing behaviors were related to state anxiety reported by the researcher, but this difference was not independent of internalizing behaviors, pointing towards a relationship via the shared variance with internalizing behaviors. Further, children with more internalizing and externalizing behaviors were less likely to participate in the actual MRI scanning procedure. Lastly, internalizing and externalizing behaviors, as well as MRI-related state anxiety were associated with worse image quality. These results underscore the potential for biases and methodological issues caused by MRI-related state anxiety in children.

## Introduction

Magnetic resonance imaging (MRI) is a medical imaging technique that has strongly enhanced our ability to understand the structure and function of the developing brain. MRI is capable of obtaining high spatial resolution images of the brain without the use of ionizing radiation, in contrast to other imaging techniques, such as Computer Tomography or Positron Emission Tomography. However, several features of the MRI procedure can be anxiety provoking. The MR gradients ramping on and off are associated with loud noises (ranging from 85 to 110 dB) (1). Moreover, those undergoing MRI are required to lie still for an extended period of time in the confined space of an MRI bore. Hence, it is common for many people to feel anxious before and during an MRI procedure.

While there is very little research on anxiety in children and adolescents undergoing MRI, in a single study by Westra et al., (2), about 50% of 5-to-12-year-old children with non-acute medical conditions (*n* = 54) undergoing diagnostic MRIs experienced anxiety and discomfort. Moreover, two studies have found that around 30% of pediatric patients experienced anxiety during an MRI procedure (3, 4). However, the anxiety may not be specifically related to the MRI, but rather the procedure, as Jaite et al. (5) recently found that undergoing an MRI did not induce more anxiety than EEG procedures in children and adolescents (aged 7–17). Trait anxiety characteristics may also play a role, as Haddad et al. (6) found that a group of clinically and sub-clinically anxious adolescents (aged 12–18) experienced higher levels of anxiety during an MRI for research purposes compared to a non-anxious group. However, this difference was attenuated by the time they returned home after the scan. Therefore, the authors concluded that MRI-related anxiety is temporary and MRI research is acceptable to adolescents, including those who are clinically anxious.

Children with MRI-related state anxiety, that do not need to undergo MRI for diagnostic purposes, may be more likely to refuse to participate in MRI related research or to be withdrawn from the procedure by researchers because of ethical concerns, as the procedure may exceed the “minimal-risk standard” (7). Anxious children and those with other behavioral problems may also be more likely to move during image acquisition, leading to motion artifacts and thus lower image quality by which they potentially introduce information bias. This means that the MRI data of children with MRI-related state anxiety, and those with internalizing and externalizing behaviors, may be more likely to be of insufficient quality for image processing. In general, up to 90% of artifacts in functional MRI can be attributed to subject movement (8). Moreover, motion artifacts, even minor movements, can impair diagnostic quality of MR examinations (9, 10). To our knowledge, whether MRI-related state anxiety has a direct impact on image quality in children has not been studied previously. There is, however, indirect evidence that pre-MRI training in children has a positive effect on image quality (11), which is presumably, at least in part, due to reductions of MRI-related anxiety.

For behavioral research, selection effects based on behavioral characteristics of the participants can have important consequences. There is evidence that, in adults who are undergoing MRI-scanning, trait anxiety is positively associated with MRI-related state anxiety (12). This could lead to selection bias, where children with the highest levels of internalizing behavior do not participate. A similar mechanism can hold for externalizing behavior as well. This bias is potentially even larger if children with the highest levels of behavioral problems are excluded from analyses. While no prior studies have assessed the relationship between state anxiety prior to and during MRI-scanning and externalizing behaviors, there are neuroimaging studies that have included non-response analyses pointing towards lower participation in those with more internalizing as well as broader problem behaviors (13, 14). In general, the relation between internalizing and externalizing behavior and MRI-related state anxiety has been understudied, but is likely to contribute to selection bias.

The current study has three aims. First, we investigate the relationship between internalizing and externalizing behavior, as measured with the Child Behavior Checklist (CBCL), and children’s MRI-related state anxiety. Second, we assess the relationship between internalizing and externalizing behavior, and MRI research participation. Lastly, we study the effects of internalizing and externalizing behavior as well as MRI-related state anxiety on image quality in children.

## Methods

### Participants

A detailed overview of the Generation R Study design and population is described previously by Jaddoe et al. (15) and a detailed overview of the neuroimaging component of the study is provided by White et al. (16). In short, the Generation R Study is an ongoing large prospective population-based cohort study with multiple waves of data collection, conducted in Rotterdam, the Netherlands. The study was approved by the Medical Ethical Committee and has been performed in accordance with the 1964 Declaration of Helsinki and its later amendments. A total of 1,932 6-10-year-old children who were invited for the first neuroimaging wave of the Generation R Study (16) were included in this study. Of those invited, 690 did not participate, due to several reasons, including inability to contact the participants, the child or the parent chose not to participate in the neuroimaging component of the study, or children could not participate due to contraindications for MRI-scanning (16). Exclusion criteria for the first neuroimaging wave included contraindications for the MRI procedure (i.e., pacemaker or ferrous metal implants), severe motor or sensory disorders (deafness or blindness), neurological disorders (i.e., seizures or tuberous sclerosis), moderate to severe head injuries with loss of consciousness, and claustrophobia.

### Procedure

The study visit consisted of a mock MRI session, followed by an actual MRI session. The mock scanner simulates the most important aspects of the actual scanning session, including the feeling of being within the MR bore, wearing headphones that play recorded gradient sounds, and the ability to watch a forward-projected film via a mirror positioned on the head coil. In this way, children could become accustomed to the scanning environment and were offered the opportunity to opt out of the procedure before going to the actual MRI scanner. Immediately after the mock scanning session, children were retrospectively asked to rate their levels of anxiety during the mock scanning procedure on a visual analog scales (VAS) with six emoji faces. If children responded that they were too scared (i.e. the sixth emoji face), they did not proceed to the actual MRI scanning session. The parents and researcher also rated children’s anxiety using the same VAS. Similarly, if either the parent or the researcher felt that the child was too scared, the child also did not proceed to the actual MRI session. Those who were comfortable undergoing the MRI-scanning procedure were scanned following the mock scanning session. Actual MRI scans were obtained from 1,070 children (86.2%).

### Magnetic Resonance Imaging

MR images were obtained with a GE Discovery MR750 3-T scanner (General Electric, Milwaukee, USA) using an 8-channel head coil for signal reception. A whole-brain high-resolution T1 inversion recovery fast spoiled gradient recalled (IR-FSPGR) sequence was obtained, with a total scan time of 5 minutes and 40 seconds. The scan parameters were: TR = 10.3 ms, TE = 4.2 ms, TI = 350ms, flip angle = 16°, matrix = 256×256, slice thickness = 0.9 mm, and in plane resolution = 0.9 × 0.9 mm.

### Measures

#### Child Behavior Checklist

During the assessment wave when children were between five and eight years of age, the CBCL 1.5–5 (17) was used to assess internalizing and externalizing behaviors in children. We used the preschool CBCL, because many children were aged younger than six years at the time of the assessment and because older-age versions are partly unsuitable for these children, as they include questions on, for example, substance use. Moreover, to enhance comparability of data between all children, the use of one version of the CBCL was preferred. The CBCL was completed by the primary caregiver, who was the mother in 93.5% of the cases. The CBCL consists of 99 items with which child behavior is rated using a three-point Likert scale (0 = not true, 1 = somewhat true, 2 = very true). Summary scores were computed for both the internalizing and externalizing scale, with higher scores indicating more problems. The CBCL has been shown the have good reliability and validity and is widely used internationally (17).

#### Visual Analog Scale

At three separate times during the neuroimaging study visit, the children were asked to indicate their levels of anxiety by using a VAS with six emoji faces (coded as 0 – 5). This was first asked before the mock scanning session. Immediately after the mock scanning session and immediately after the actual MRI scanning session, children were asked to retrospectively rate their anxiety levels during the (mock) MRI session. The parent (one parent per child) and researcher also rated the level of anxiety of the child at these three time points (16). We have utilized a multi-informant approach, because all informants can contribute unique and valuable information (18). The VAS is similar to that developed by Durston et al. (19).

#### Image quality

The automated quality assessment rating was based on quantifying the blurring of the edge spread function at the border of the head that is associated with head movement during scanning (10). This algorithm provides a fine detailed measure with a Gaussian distribution of motion artifacts during scanning. A detailed description of this automated quality assessment tool is described in White et al., 2018 (10).

### Statistical analyses

All analyses were performed using the Statistical Package for the Social Sciences (version 25, IBM Corp., Armonk, NY) for Windows.

#### MRI-related anxiety

Our first goal was to test the hypothesis that internalizing and externalizing behaviors (CBCL) were associated with children’s state anxiety (VAS) during an (actual) MRI procedure. We used a linear mixed model analysis to determine whether there was a relationship between internalizing behavior (CBCL) and MRI-related state anxiety (VAS) reported by the child, parent, or researcher. The VAS scores were entered as three independent variables (child, parent and researcher), with three time points each (before the mock MRI, during the mock MRI, and during the actual MRI). In the first model, we corrected for age and sex. In the second model, we corrected for age, sex, and externalizing behavior. We repeated this mixed model analysis to determine whether there was a relationship between externalizing behavior and MRI-related state anxiety. In this analysis, we corrected for age, sex, and internalizing behavior in the second model.

#### MRI participation

We tested the hypothesis that child internalizing and externalizing behaviors impact MRI research participation. Logistic regression was used to assess whether internalizing and externalizing behaviors were related to MRI participation. First, internalizing and externalizing behaviors were entered as independent variables in separate models, with MRI participation as dependent variable. These analyses were corrected for age and sex. Second, internalizing and externalizing behaviors were entered in the same model, while additionally correcting for age and sex, to assess to what extent they were independently related to MRI participation. Since child anxiety levels before and during the mock scanner were integral to the decision to continue with the actual MRI procedure, we chose not to investigate the role of MRI-related state anxiety as a predictor of MRI participation.

#### MRI image quality

We investigated whether internalizing and externalizing behaviors, as well as state anxiety during the actual MRI were related to MRI image quality. We performed an overall analysis by means of mixed modelling. The internalizing and externalizing CBCL scores were entered in the model together with the VAS scores during the MRI reported by the child, parent, and researcher, as independent variables, VAS scores were added as three levels of one factor. The analysis was corrected for age and sex. Automatically assessed MRI image quality was used as the outcome.

#### Multiple testing correction

We performed multiple testing correction on the second models of our main analyses, which are the two mixed models to investigate the relation between internalizing and externalizing behaviors, and MRI related state anxiety, the two logistic regression models to investigate the relation between internalizing and externalizing behaviors and MRI participation, and the mixed model investigating the relation between internalizing and externalizing behaviors and MRI related state anxiety, and MRI image quality, using the FDR-Benjamini Hochberg procedure for a total of 11 tests (20).

## Results

### Descriptive statistics

The mean age of children who participated in the current study (*n* = 1,241) was 6.13 years (*SD* = 0.46). A fairly equal amount of boys (53.2%) and girls (46.8%) participated in the study. Most mothers had high (45.4%) or medium (47.1%) educational levels, compared to low educational levels (7.5%). The majority of participants (59.5%) had a monthly household income >€2000; 19.9% had a household income of €1200-2000, and 15.3% had a household income <€1200. Characteristics of children who were invited, but did not participate in the current study were similar to characteristics of participants, except for maternal educational level, which was significantly higher for children who did participate compared to those who did not.

Mean summary scores for internalizing and externalizing behaviors were 8.30 (SD: 7.52) and 10.29 (SD: 8.23), respectively. Child-, parent-, and researcher-reported mean state anxiety levels of children during the actual MRI are 0.71, 0.97, and 0.90, respectively (see Table 1). Moderate to strong levels of correspondence between informants were found for MRI-related anxiety before mock scanning, during mock scanning, and during MRI (see supplementary materials). Additionally, a large significant correlation of 0.73 between internalizing and externalizing behaviors was observed.

**Table 1:**
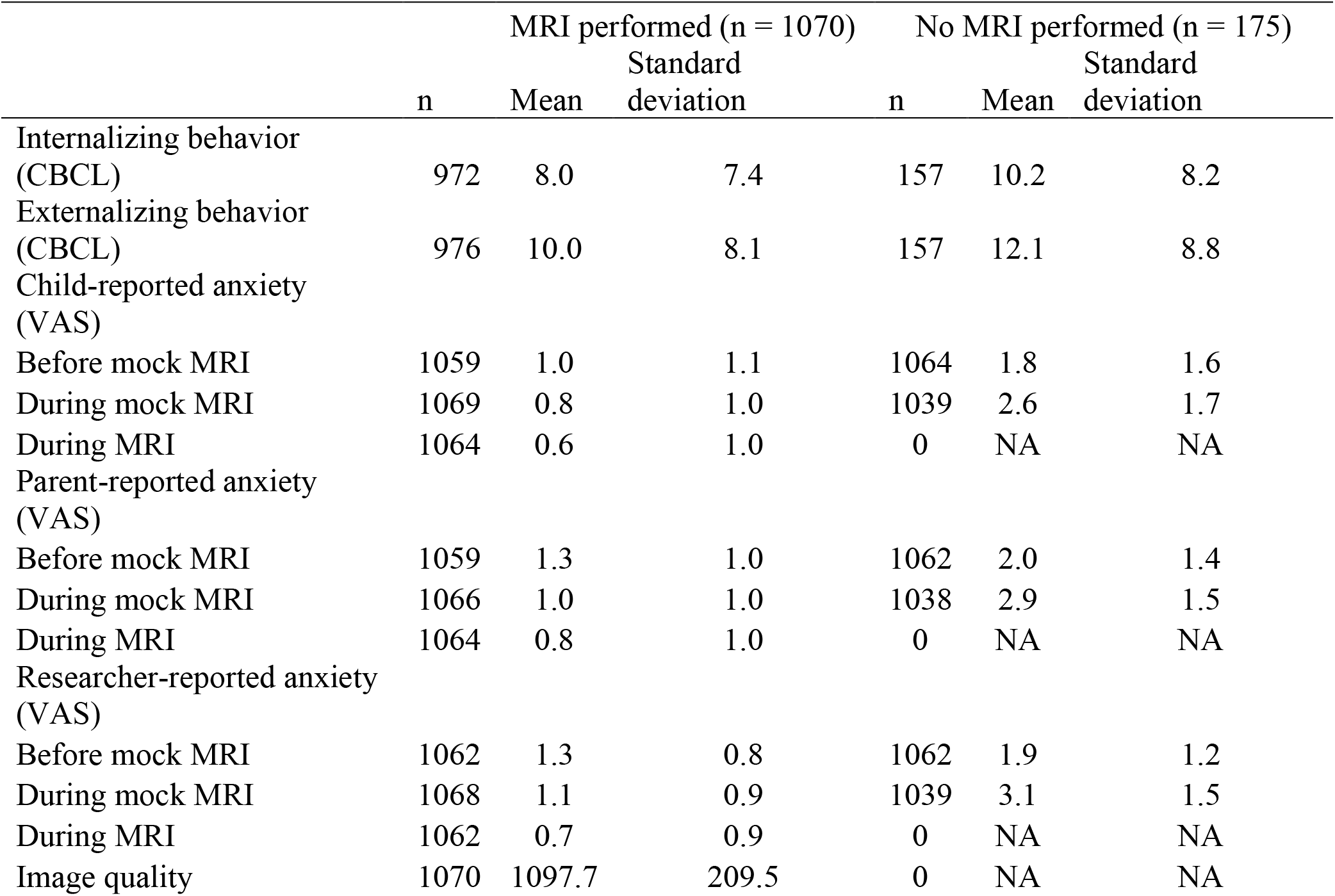
Descriptive statistics

|  | MRI performed (n = 1070) |  |  | No MRI performed (n = 175) |  |  |
| --- | --- | --- | --- | --- | --- | --- |
|  | n | Mean | Standard deviation | n | Mean | Standard deviation |
| Internalizing behavior (CBCL) | 972 | 8.0 | 7.4 | 157 | 10.2 | 8.2 |
| Externalizing behavior (CBCL) | 976 | 10.0 | 8.1 | 157 | 12.1 | 8.8 |
| Child-reported anxiety (VAS) |  |  |  |  |  |  |
| Before mock MRI | 1059 | 1.0 | 1.1 | 1064 | 1.8 | 1.6 |
| During mock MRI | 1069 | 0.8 | 1.0 | 1039 | 2.6 | 1.7 |
| During MRI | 1064 | 0.6 | 1.0 | 0 | NA | NA |
| Parent-reported anxiety (VAS) |  |  |  |  |  |  |
| Before mock MRI | 1059 | 1.3 | 1.0 | 1062 | 2.0 | 1.4 |
| During mock MRI | 1066 | 1.0 | 1.0 | 1038 | 2.9 | 1.5 |
| During MRI | 1064 | 0.8 | 1.0 | 0 | NA | NA |
| Researcher-reported anxiety (VAS) |  |  |  |  |  |  |
| Before mock MRI | 1062 | 1.3 | 0.8 | 1062 | 1.9 | 1.2 |
| During mock MRI | 1068 | 1.1 | 0.9 | 1039 | 3.1 | 1.5 |
| During MRI | 1062 | 0.7 | 0.9 | 0 | NA | NA |
| Image quality | 1070 | 1097.7 | 209.5 | 0 | NA | NA |

**Table 2:** Association between internalizing and externalizing behavior, and MRI participation (reference group: participated in MRI scanning). Model 1: corrected for age and sex, Model 2: internalizing and externalizing behavior in one model, corrected for age and sex.

| Behavior | Model | Odds Ratio<br>MRI<br>participation | 95%<br>Confidence<br>Interval | p-value |
| --- | --- | --- | --- | --- |
| Internalizing | 1 | 1.03 | 1.01-1.06 | 0.001 |
|  | 2 | 1.03 | 1.00-1.06 | 0.09 |
| Externalizing | 1 | 1.03 | 1.01-1.05 | 0.004 |
|  | 2 | 1.01 | 0.98-1.04 | 0.49 |

### MRI-related anxiety

In the overall mixed model analysis, when correcting for age and sex, internalizing behaviors, rated on average 1.82 years (SD = 0.94) before the MRI procedure, were significantly associated with MRI-related state anxiety across all three time points as reported by the child (*F* (5, 3172) = 2.565, *p* = 0.025), the parent (*F* (5, 3172) = 6.773, *p* < 0.001) and the researcher (*F* (5, 3172) = 4.300, *p* = 0.001). When correcting for age, sex, and externalizing behaviors, internalizing behaviors were significantly associated with MRI-related state anxiety across all three time points as reported by the parent (*F* (5, 3165) = 6.652, *p* < 0.001) and the researcher (*F* (5, 3165) = 2.896, *p* = 0.013), but not as reported by the child (*F* (5, 3165) = 1.400, *p* = 0.221). Both the relationship between parent- and researcher-reported state anxiety, and internalizing behaviors, remained statistically significant after multiple testing correction.

In the overall mixed model for externalizing behavior, results were more ambiguous. Over all three time points combined, when correcting for age and sex, externalizing behavior was not associated with child-reported (*F* (5, 3181) = 1.565, *p* = 0.166) or parent-reported MRI-related state anxiety of the child (*F* (5, 3181) = 2.061, *p* = 0.067). Externalizing behaviors were significantly associated with higher researcher-reported state anxiety of the child (*F* (5, 3172) = 2.346, *p* = 0.039), correcting for age and sex. After additional correction for internalizing behavior, externalizing behavior was not associated with MRI-related state anxiety, when reported by the child, parent, or researcher (*F* (5, 3165) = 0.309, *p* = 0.908, *F* (5, 3165) = 1.805, *p* = 0.108, *F* (5, 3165) = 1.068, *p* = 0.376, respectively).

### MRI participation

Logistic regression analyses revealed that more internalizing (odds ratio = 1.03; 95% CI = 1.01, 1.06) and externalizing (odds ratio = 1.03; 95% CI = 1.01, 1.05) behaviors were related to a lower probability of MRI participation when correcting for age and sex only. However, when both internalizing and externalizing behaviors were entered in the same model, these associations did not reach statistical significance (Table 4).

### Image quality

The mixed model analysis indicated that more internalizing (*F* (1, 2880) = 5.823, *p* = 0.016) and externalizing behavior (*F* (1, 2880) = 10.716, *p* = 0.001) was associated with poorer MRI image quality. In addition, more MRI-related state anxiety (during the actual MRI procedure) was associated with poorer MRI image quality (*F* (1, 2880) = 21.223, *p* < 0.001). All results remained statistically significant after multiple testing correction.

## Discussion

This study was aimed at investigating the relationship between children’s MRI-related state anxiety and internalizing and externalizing symptoms, as a source of potential bias for participation as well as image quality, in a large population sample of school-aged children. Internalizing behavior was significantly associated with MRI-related state anxiety, as reported by the child, parent, and researcher, when corrected for age and sex. After additional correction for externalizing behavior, this association was still significant for parent- and researcher-reported anxiety, but not for child-reported anxiety. Regarding externalizing behaviors, only the relationship with researcher reported state anxiety reached statistical significance. However, this relationship was no longer apparent after additional correction for internalizing behaviors. Furthermore, our results indicate that children with more internalizing or externalizing behaviors are less likely to participate in an MRI examination. Finally, we found evidence that internalizing and externalizing behaviors, as well as higher levels of state anxiety during the MRI procedure are associated with poorer image quality.

Where numerous earlier studies have focused on reducing state anxiety prior to MRI-scanning to improve image quality (21-24) and guides have been written for MRI-technologists on how to reduce state anxiety in people undergoing MRI-scanning (25), no prior studies have assessed whether state anxiety was related to behavioral traits, such as internalizing and externalizing behavior. Our results indicate that internalizing and externalizing behaviors are both associated with state anxiety and that internalizing behaviors are associated with state anxiety, independent of externalizing behaviors. For externalizing behaviors, associations were absent when correcting for internalizing behaviors, indicating that the relationship between externalizing behaviors and state anxiety is explained by the shared variance between internalizing and externalizing behaviors.

These findings suggest the importance of a careful assessment of children’s behavioral characteristics prior to MRI-scanning. Since children with more internalizing and externalizing symptoms also have higher state anxiety, those children would likely benefit most from interventions reducing state anxiety. The result would be better retention of research participants and better quality data. Tailored interventions for those with more trait problems could be implemented to assess the extent to which state anxiety can be reduced to increase MRI participation and image quality. Our findings on the relation between state anxiety and internalizing and externalizing behavior are important, as this relationship can potentially impact results for MRI studies assessing internalizing and externalizing behaviors. Most notably, this can impact functional MRI (fMRI) studies, since these studies assess subtle differences in brain activation. Because of the overlap in brain networks activated by state anxiety, trait anxiety, and pathological anxiety (26-28), MRI-related state anxiety may act as a moderating factor between internalizing behavior and functional MRI (fMRI) responses. This could implicate that state anxiety could exacerbate differences in brain activation between children with internalizing behavior, such as anxiety and depression, and children without such behavior. Second, MRI-related state anxiety could also be of concern for studies in children with externalizing behavior, because fMRI signals in certain brain regions could be attributed to the externalizing behavior under study, while in reality, the signals are partly related to state anxiety induced by the MRI procedure. An additional problem is that when MRI-related state anxiety reduces over time, across multiple scanning sessions, the differences in fMRI outcomes between sessions could be wrongly interpreted, for example as a treatment effect (29). Thus, the next crucial step that needs to be addressed in future work is to what extent state anxiety confounds fMRI studies on internalizing and externalizing behaviors.

Children who did not participate in the actual MRI examination because they were too anxious at that moment, had more internalizing and externalizing behaviors than children who did participate. These differences were present despite the fact that the overall dropout rate among those who wanted to participate in our study was low (13.8% of 1,241 children), which is possibly due to our use of a mock MRI scanner. Our findings indicate the potential for selection bias, which can impact all MRI research, as it may dilute the number of participants with higher levels of internalizing or externalizing symptoms included in MRI samples. In turn, this could diminish the power of studies to find relevant associations between (f)MRI features and internalizing or externalizing symptoms, and may reduce the strength of the effect of these associations. Based on these results we recommend future studies using samples in which children voluntarily underwent MRI-scanning to check whether selection bias is present in their sample.

We found that MRI quality was impacted by internalizing and externalizing behaviors, as well as by MRI-related state anxiety. Children with more internalizing and externalizing behaviors as well as those with more state anxiety were more likely to move during image acquisition, leading to lower quality images. This has important implications, because participants with poor image quality are often removed from analyses (30), but if not removed, movement during both structural and functional neuroimaging can alter quantitative brain metrics (10, 31, 32). This means that children with elevated state and trait problems, in addition to being more likely to drop out prior to the actual scanning procedure, have a higher chance to be removed from analyses, which potentially further increases the aforementioned bias in the results. The observed association is not in line with previous findings in adult samples. For example, Dantendorfer et al. (9) did not find a relationship between anxiety and image quality in adult participants. However, this discrepancy in findings may be explained by methodological differences. The authors used a dichotomous outcome for motion artifacts, which limits the power and generalizability of the study, since only 13% of the MRI sequences were unusable. In contrast, our study has much more power and we used image quality as continuous variable. Klaming et al. (33) also did not find a relationship between anxiety and motion artifacts in adult patients, but a limitation of this study includes the image analysis software with a low temporary sampling rate, which was not able to detect intra-scan movements. Another possible explanation is that the relationship between state anxiety and MRI quality is more profound in children than in adults. Compared to children, adults may have a better understanding of the MRI procedures, have developed more coping strategies to manage stressful situations, and have stronger inhibitory control which enables them to lie still during the MRI (34, 35). It is important to further investigate the relationship between MRI-related anxiety and image quality in children, because movement artifacts result in non-Gaussian, or colored, noise (e.g., reduced cortical thickness), which could increase false positive findings (10).

Taken together, these findings underscore the importance of dealing with MRI-related state anxiety. Options are to either investigate ways to statistically correct for MRI-related anxiety or to focus on reducing anxiety concerning an MRI procedure, especially in those with more internalizing and externalizing behavior. A statistical option could be, for example, to correct for state anxiety by adding this variable as covariate to analyses. However, a drawback of this approach could be overcorrection of state anxiety. On the other hand, reducing MRI-related state anxiety could possibly be achieved by (repeated) mock MRI sessions (29), educational videos (23), or habituation through virtual reality (22), although it should be noted that our study shows that even a proactive approach to reduce MRI-related anxiety via preparation with a mock MRI scanner, this did not fully remove the issue of MRI-related state anxiety. Reducing state anxiety is especially important for children who need to undergo a diagnostic MRI, as they usually do not have access to a mock scanner and reducing MRI-related anxiety could preclude the need for sedation.

## Strengths and limitations

An important strength of this study is the large sample size. Other strengths include the usage of multiple informants of child anxiety, inclusion of both a mock and an actual MRI examination, and the use of an automated quality assessment measure for the structural MRI. One aspect of the study that can be considered both a strength and a limitation is the narrow age range. The narrow age range is a strength, as it provides a homogeneous group of children where age influences are minimized. However, because we assessed specifically a rather narrow age-range of children we cannot be certain that these results generalize to other age ranges. Second, the CBCL data were collected in advance of the MRI scans, with a mean time interval between CBCL data collection and MRI scan of 1.8 years. However, CBCL scores have shown to be stable over time (36, 37). Further, in the current study we have collected data on state anxiety during the MRI scanning by asking children about their anxiety immediately after the MRI procedure. A more optimal approach would have been to ask them about their anxiety while they were still in the scanner. Children who decided not to participate due to being anxious about the MRI did not visit the MRI center, nor receive a mock scan. Thus, we may not have seen the most anxious children. Finally, the procedure introduced has two limitations. First, since MRI participation was based on state anxiety before or during mock scanning, we were not able to assess to what extent non-participation was related to internalizing and externalizing behaviors independent of state anxiety. Second, we have possibly underestimated the magnitude of the effect of MRI-related state anxiety on image quality, because children with the highest levels of state anxiety before or during the mock MRI did not proceed to the actual MRI scanning session and because children who did proceed to the actual MRI, already underwent a mock MRI that was aimed at reducing anxiety during the actual MRI. Despite this, we still found evidence that MRI-related state anxiety introduces methodological issues in MRI research.

## Conclusion

In conclusion, our study demonstrates a positive association between both internalizing and externalizing behaviors and MRI-related state anxiety in a large population sample of school-aged children. This indicates that MRI-related state anxiety can influence MRI research on internalizing and externalizing behaviors. Ultimately, this may lead to erroneous interpretation of MRI results, which is especially a concern for etiologically oriented research. Moreover, internalizing behavior, externalizing behavior, and MRI-related state anxiety impact MRI participation and image quality. It is important to investigate optimal approaches to adjust for the potential moderating or mediating effects of MRI-related state anxiety in future (f)MRI studies on psychopathology symptoms in children. Moreover, it is important to investigate interventions to reduce anxiety surrounding an MRI procedure, especially in those children with increased internalizing and externalizing behaviors.

## Data Availability

Generation R data is available to researchers and requests should be directed to the management team of the Generation R Study. Release of data is contingent upon privacy and ethical restrictions.

## Acknowledgements

This work was supported by Netherlands Organization for Health Research and Development (ZonMw) TOP [grant number 91211021]. The Generation R Study is conducted by the Erasmus Medical Center in close collaboration with the School of Law and Faculty of Social Sciences of the Erasmus University Rotterdam, the Municipal Health Service Rotterdam area, Rotterdam, the Rotterdam Homecare Foundation, Rotterdam and the Stichting Trombosedienst and Artsenlaboratorium Rijnmond (STAR-MDC), Rotterdam. We gratefully acknowledge the contribution of children and parents, general practitioners, hospitals, midwives, and pharmacies in Rotterdam. The authors would like to thank Nikita Schoemaker and Marcus Schmidt for their coordination and (technical) support within the Generation R pilot brain imaging study.

## Notes

**Funding** This work was supported by Netherlands Organization for Health Research and Development (ZonMw) TOP [grant number 91211021]; Netherlands Organization for Scientific Research (NWO) Brain and Cognition [433-09-228]; Erasmus Medical Center, Rotterdam; Erasmus University Rotterdam; Ministry of Health, Welfare and Sport; Ministry of Youth and Families

### Competing Interest Statement

The authors have declared no competing interest.

### Author Declarations

This study was approved by the Medical Ethical Committee of the Erasmus Medical Center and has been performed in accordance with the ethical standards laid down in the 1964 Declaration of Helsinki and its later amendments. All persons gave their informed consent prior to their inclusion in the study.

## References

1. Price DL, De Wilde JP, Papadaki AM, Curran JS, Kitney RI. Investigation of acoustic noise on 15 MRI scanners from 0.2 T to 3 T. J Magn Reson Imaging. 2001;13(2):288–93.

2. Westra AE, Zegers MP, Sukhai RN, Kaptein AA, Holscher HC, Ballieux BE, et al. Discomfort in children undergoing unsedated MRI. Eur J Pediatr. 2011;170(6):771–7.

3. Marshall SP, Smith MS, Weinberger E. Perceived anxiety of pediatric patients to magnetic resonance. Clinical pediatrics. 1995;34(1):59–60.

4. Tyc VL, Fairclough D, Fletcher B, Leigh L, Mulhern RK. Children’s distress during magnetic resonance imaging procedures. Children’s Health Care. 1995;24(1):5–19.

5. Jaite C, Kappel V, Napp A, Sommer M, Diederichs G, Weschke B, et al. A comparison study of anxiety in children undergoing brain MRI vs adults undergoing brain MRI vs children undergoing an electroencephalogram. PloS one. 2019;14(3):e0211552.

6. Haddad ADM, Platt B, James AC, Lau JYF. Anxious and non-anxious adolescents’ experiences of non-clinical magnetic resonance imaging research. Child Psychiatry & Human Development. 2013;44(4):556–60.

7. Schmidt MH, Marshall J, Downie J, Hadskis MR. Pediatric magnetic resonance research and the minimal-risk standard. IRB-Ethics and Human Research. 2011;33(5):1.

8. Friston KJ, Williams S, Howard R, Frackowiak RS, Turner R. Movement-related effects in fMRI time-series. Magn Reson Med. 1996;35(3):346–55.

9. Dantendorfer K, Amering M, Bankier A, Helbich T, Prayer D, Youssefzadeh S, et al. A study of the effects of patient anxiety, perceptions and equipment on motion artifacts in magnetic resonance imaging. Magn Reson Imaging. 1997;15(3):301–6.

10. White T, Jansen PR, Muetzel RL, Sudre G, El Marroun H, Tiemeier H, et al. Automated quality assessment of structural magnetic resonance images in children: Comparison with visual inspection and surface based reconstruction. Human brain mapping. 2018;39(3):1218–31.

11. Viggiano MP, Giganti F, Rossi A, Di Feo D, Vagnoli L, Calcagno G, et al. Impact of psychological interventions on reducing anxiety, fear and the need for sedation in children undergoing magnetic resonance imaging. Pediatric reports. 2015;7(1).

12. Thu HS, Stutzman SE, Supnet C, Olson DM. Factors associated with increased anxiety in the MRI waiting room. Journal of Radiology Nursing. 2015;34(3):170–4.

13. Radoman M, Phan KL, Gorka SM. Neural correlates of predictable and unpredictable threat in internalizing psychopathology. Neuroscience letters. 2019;701:193–201.

14. Muetzel RL, Mulder RH, Lamballais S, Cortes Hidalgo AP, Jansen P, Güroğlu B, et al. Frequent bullying involvement and brain morphology in children. Frontiers in psychiatry. 2019;10:696.

15. Jaddoe VWV, Mackenbach JP, Moll HA, Steegers EAP, Tiemeier H, Verhulst FC, et al. The Generation R Study: design and cohort profile. European journal of epidemiology. 2006;21(6):475.

16. White T, El Marroun H, Nijs I, Schmidt M, van der Lugt A, Wielopolki PA, et al. Pediatric population-based neuroimaging and the Generation R Study: the intersection of developmental neuroscience and epidemiology. European journal of epidemiology. 2013;28(1):99–111.

17. Achenbach TM, Rescorla LA. Manual for the ASEBA preschool forms & profiles. Burlington: University of Vermont, Research Center for Children, Youth, & Families; 2000.

18. De Los Reyes A, Thomas SA, Goodman KL, Kundey SMA. Principles underlying the use of multiple informants’ reports. Annual review of clinical psychology. 2013;9:123–49.

19. Durston S, Nederveen H, van Dijk S, van Belle J, de Zeeuw P, Langen M, et al. Magnetic resonance simulation is effective in reducing anxiety related to magnetic resonance scanning in children. Journal of the American Academy of Child & Adolescent Psychiatry. 2009;2(48):206–7.

20. Hochberg Y. A sharper Bonferroni procedure for multiple tests of significance. Biometrika. 1988;75(4):800–2.

21. Ajam AA, Tahir S, Makary MS, Longworth S, Lang EV, Krishna NG, et al. Communication and team interactions to improve patient experiences, quality of care, and throughput in MRI. Topics in Magnetic Resonance Imaging. 2020;29(3):131–4.

22. Ashmore J, Di Pietro J, Williams K, Stokes E, Symons A, Smith M, et al. A Free Virtual Reality Experience to Prepare Pediatric Patients for Magnetic Resonance Imaging: Cross-Sectional Questionnaire Study. JMIR Pediatrics and Parenting. 2019;2(1):e11684.

23. Szeszak S, Man R, Love A, Langmack G, Wharrad H, Dineen RA. Animated educational video to prepare children for MRI without sedation: evaluation of the appeal and value. Pediatric radiology. 2016;46(12):1744–50.

24. Grey SJ, Price G, Mathews A. Reduction of anxiety during MR imaging: a controlled trial. Magnetic resonance imaging. 2000;18(3):351–5.

25. Stogiannos N. Reducing patient’s psychological stress. A guide for MR technologists. Hellenic Journal of Radiology. 2019;4(1).

26. Takagi Y, Sakai Y, Abe Y, Nishida S, Harrison BJ, Martinez-Zalacain I, et al. A common brain network among state, trait, and pathological anxiety from whole-brain functional connectivity. Neuroimage. 2018;172:506–16.

27. Deckersbach T, Dougherty DD, Rauch SL. Functional imaging of mood and anxiety disorders. Journal of Neuroimaging. 2006;16(1):1–10.

28. Tian X, Wei D, Du X, Wang K, Yang J, Liu W, et al. Assessment of trait anxiety and prediction of changes in state anxiety using functional brain imaging: A test-retest study. Neuroimage. 2016;133:408–16.

29. Chapman HA, Bernier D, Rusak B. MRI-related anxiety levels change within and between repeated scanning sessions. Psychiatry Research: Neuroimaging. 2010;182(2):160–4.

30. Andre QR, Geeraert BL, Lebel C. Brain structure and internalizing and externalizing behavior in typically developing children and adolescents. Brain Structure and Function. 2019:1–10.

31. Power JD, Mitra A, Laumann TO, Snyder AZ, Schlaggar BL, Petersen SE. Methods to detect, characterize, and remove motion artifact in resting state fMRI. Neuroimage. 2014;84:320–41.

32. Satterthwaite TD, Wolf DH, Loughead J, Ruparel K, Elliott MA, Hakonarson H, et al. Impact of in-scanner head motion on multiple measures of functional connectivity: relevance for studies of neurodevelopment in youth. Neuroimage. 2012;60(1):623–32.

33. Klaming L, van Minde D, Weda H, Nielsen T, Duijm LE. The Relation Between Anticipatory Anxiety and Movement During an MR Examination. Acad Radiol. 2015;22(12):1571–8.

34. Rosenberg DR, Sweeney JA, Gillen JS, Kim J, Varanelli MJ, O’Hearn KM, et al. Magnetic resonance imaging of children without sedation: preparation with simulation. Journal of the American Academy of Child & Adolescent Psychiatry. 1997;36(6):853–9.

35. Greenberg SB, Faerber EN, Radke JL, Aspinall CL, Adams RC, Mercer-Wilson DD. Sedation of difficult-to-sedate children undergoing MR imaging: value of thioridazine as an adjunct to chloral hydrate. AJR American journal of roentgenology. 1994;163(1):165–8.

36. Stanger C, MacDonald VV, McConaughy SH, Achenbach TM. Predictors of cross-informant syndromes among children and youths referred for mental health services. Journal of Abnormal Child Psychology. 1996;24(5):597–614.

37. Verhulst FC, Van der Ende J. Six-year stability of parent-reported problem behavior in an epidemiological sample. Journal of abnormal child psychology. 1992;20(6):595–610.

