## Supplementary Table 1 for "Internalizing and externalizing behaviors in school-aged children are related to state anxiety during magnetic resonance imaging"

Supplementary Table 1: Correlations between child-reported, parent-reported, and researcher-reported state anxiety (VAS) before the mock MRI, during the mock MRI, and during the actual MRI.

| | child - before mock | child – during mock | child - during MRI | parent - before mock | parent - during mock | parent - during MRI | researcher - before mock | researcher - during mock | researcher - during MRI |
|---|---|---|---|---|---|---|---|---|---|
| child - before mock | 1 | .340** | .194** | .515** | .351** | .171** | .587** | .371** | .170** |
| child - during mock | .340** | 1 | .324** | .293** | .664** | .340** | .332** | .741** | .356** |
| child - during MRI | .194** | .324** | 1 | .161** | .266** | .696** | .172** | .315** | .749** |
| parent - before mock | .515** | .293** | .161** | 1 | .487** | .218** | .563** | .370** | .157** |
| parent - during mock | .351** | .664** | .266** | .487** | 1 | .418** | .395** | .741** | .348** |
| parent - during MRI | .171** | .340** | .696** | .218** | .418** | 1 | .188** | .354** | .769** |
| researcher - before mock | .587** | .332** | .172** | .563** | .395** | .188** | 1 | .484** | .225** |
| researcher - during mock | .371** | .741** | .315** | .370** | .741** | .354** | .484** | 1 | .463** |
| researcher - during MRI | .170** | .356** | .749** | .157** | .348** | .769** | .225** | .463** | 1 |

**. Correlation is significant at the 0.01 level (2-tailed).
